# Impact of comorbidities on COVID-19 outcome

**DOI:** 10.1101/2020.11.28.20240267

**Authors:** Eman M khedr, Enas Daef, Aliae Mohamed-Hussein, Ehab F Mostafa, Mohamed zein, Sahar M Hassany, Hanan Galal, Shimaa Abbas Hassan, Islam Galal, Amro A. Zarzour, Helal F Hetta, Hebatallah M. Hassan, Mariam Taher Amin, Maiada k Hashem

## Abstract

**Background and aims:** The coronavirus disease 19 (COVID-19) pandemic has spread rapidly around the globe with considerable morbidity and mortality. Coexistence of comorbidities with COVID-19 have consistently been reported as risk factors for unfavorable prognosis. We aim at this study to evaluate the impact of comorbidities in COVID-19 patients on the outcome and determine predictors of prolonged hospital stay, requisite for ICU admission or decease.

**Methods:** Four hundreds and thirty nine adult patients who are admitted through (June and July 2020) in Assiut and Aswan University Hospitals were included in the study. All participants were diagnosed with COVID-19 according to Egyptian Ministry of Health guidance as definite case or Probable case. Detection of SARS-CoV-2 RNA was done by (TaqMan™ 2019-nCoV Control Kit v1 (Cat. No. A47532) supplied by QIAGEN, Germany on the Applied Biosystem 7500 Fast RT PCR System, USA.

**Results:** Patients with comorbidities represented 61.7% of all cases. Constitutional symptoms especially myalgia and LRT symptoms such as dyspnea were significantly higher in patients with comorbidities (P < 0.05). Patients with comorbidities had significantly worse laboratory parameters. ICU admission was higher in patients with comorbidities (35.8%). Among different comorbidities 45.4% of CVD cases were admitted in ICU followed by DM cases (40.8%). Also, patients with comorbidities needed invasive mechanical ventilation more than those without comorbidity (31 vs. 10.7%, P<0.001). Significant lower frequency of recovery was found in COVID-19 patients with comorbidities (59% vs. 81%, P<0.001) and death rate was significantly higher in cases with comorbidities (P< 0.001). The survival rates in cases with pre-existing CVD and neurological diseases were lower than those without disease (P<0.002 and 0.001 respectively).

**Conclusion:** Association of cardiovascular comorbid conditions including hypertension or neurological diseases together with COVID-19 infections carries higher risks of mortality. However, other comorbidities such as diabetes mellitus, chronic pulmonary or kidney diseases may also contribute to increased COVID-19 severity.

## Introduction

COVID-19 is a novel emerging, rapidly propagating illness that is overwhelming most of resources of efficient health-care systems, and numerous hospitals, globally, are presently suffering a lack of ICU beds for critically ill COVID-19 pneumonic patients. Till now more than 26 million cases and 850 thousand deceases were recorded worldwide according to WHO(1). A risk stratification based on clinical, radiological and laboratory parameters seems necessary in order to better identify those patients who may need hospital and/or ICU admission.

Generally, one of the ultimate alarming clinical considerations is the presence of comorbidities. Comorbidities are associated with worse health outcome, more complex clinical management, and increased health care cost. We should seek to provide the best local solutions in conjunction with the recent national guidelines to continue the proper management of patients while ensuring proper resource (2). Chronic disorders participate in numerous topographies with communicable diseases, for instance the proinflammatory state, and the attenuation of the innate immune response which may possibly be allied etiologically to its pathogenesis (3). Moreover, upregulation of angiotensin-converting enzyme in patients with chronic diseases are increasing the susceptibility to SARS-CoV-2 infection and the risk of disease aggravation (4).

A recent meta-analysis reported that underlying disease, including hypertension, diabetes mellitus, respiratory and cardiovascular disease (5), as well as obesity(6), may be risk factors for adverse outcomes. So further studies of comorbidities as a risk of fatality at different communities are required.(7).

**The aim** of this study is to evaluate the impact of comorbidities in COVID-19 patients on the outcome and determine predictors of prolonged hospital stay, requisite for ICU admission or decease.

### Patients and methods

#### Study design and population

This is a retrospective observational cohort study conducted in two major health-care centers in Upper Egypt. First one was Assiut University Hospitals which involve 7 specialized hospitals and second one was Aswan University Hospital. Both were designated to diagnose and treat moderate and severe cases of COVID-19.

All adult patients admitted through (June and July 2020) were included in the study.

#### Diagnosis and Evidence of COVID-19

All participants were diagnosed with COVID-19 according to Egyptian Ministry of Health guidance. Evidence of SARS-CoV-2 infection was defined as

1. Cases with definite COVID-19 if patients came with clinical symptoms of infection and PCR of respiratory samples (eg, nasal or throat swab) was positive.
2. Cases with probable COVID-19 if clinical symptoms of infection and chest CT was consistent with COVID-19 plus one or 2 laboratory investigations were positive) (lymphopenia, high serum ferretin and D-Dimer Level) but PCR was negative or not done.

After reviewing of records any patient less than 18 years or with missed clinical outcomes status was excluded from the study.

#### PCR diagnosis has been done as the following

##### Specimens collection

For initial diagnostic testing for SARS-CoV-2, CDC recommends collecting and testing an upper respiratory specimen. The following specimens were collected by a health-care provider from cases with suspected novel coronavirus infection: A nasopharyngeal (NP) or oropharyngeal (OP).

Swab specimens were collected using swabs with a synthetic tip, and an aluminum or plastic shaft. Swabs were placed immediately into sterile tubes containing 2-3 mL of viral transport media. The samples were stored at 2-8°C up to 72 hours.

1. **RNA extraction of SARS-CoV-2** was done using (QIAamp Viral RNA Mini Kit Catalog no. 52904 supplied by QIAGEN, Germany). Sample preparation using QIAcube instruments follows the same steps as the manual procedure (i.e., lyse, bind, wash and elute).
2. **Pathogen detection of SARS-CoV-2 RNA** was done by (TaqMan™ 2019-nCoV Control Kit v1 (Cat. No. A47532) supplied by QIAGEN, Germany) on the Applied Biosystem 7500 Fast RT PCR System, USA. It amplified and detected three viral genomic regions, reducing the risk of false negatives including the N protein (nucleocapsid gene), S protein (Spike gene), and open reading frame-1ab (ORF1ab) genes. Applied Biosystems™ TaqMan™ 2019-nCoV Control Kit v1 (Cat. No. A47533) is a synthetic positive control that contains target sequences for each of the assays included in the TaqMan™ 2019-nCoV Assay Kit v1 (Cat. No. A47532).

#### Data collection

Clinical records and laboratory data were reviewed by the investigators in each study site and the following data were extracted for analysis:

1. ***Demographic and clinical data***: age, gender, presenting symptoms, comorbidities, and outcomes.
2. ***Laboratory investigations:*** complete blood picture, liver function tests, kidney function tests, D-dimer, serum ferritin and C-reactive protein (CRP).
3. ***Chest CT findings***.
4. ***Clinical outcomes***: complete recovery, need for ICU admission, and death.

*Classification of the patients:* According to the presence or absence of preexisiting comorbidities which determined based on patient’s self-report on admission if diagnosed with any chronic illness on the last 10 years then comorbidities were classified based on the number (single, two and three or more); the patients were classified into

a. **Group I: COVID-19 patients with** at least one of the comorbidities (diabetes mellitus, cardiovascular diseases including hypertension, chronic pulmonary disease, chronic liver disease, chronic kidney disease, neurological disease, autoimmune disease, endocrinal disease and any other disease or risk factor).
b. **Group II: COVID-19 patients without any of comorbidities**

#### Ethical approval

The research protocol was approved via the Ethical Review Committee of Assiut Faculty of Medicine before starting of the study ***(IRB no: 17300434)***. Patients identifying information were concealed and each patient assigned for a code to insure privacy and confidentiality of the data. Written informed consent was obtained from the each patient or from first degree relatives.

### Statistical analysis

All statistical analyses were performed using IBM SPSS Statistics version 20 (SPSS Inc., Chicago, IL, USA). Categorical data were presented as frequencies and percentages, while Chi-square tests were used for comparisons between groups. Continuous data were reported as means ± standard deviations and tested for normality using the Shapiro-Wilkes test. Where continuous data were normally distributed, the Student’s T-test was used for comparisons between groups; where data were non-normally distributed, the Mann-Whitney test was used. For survival analysis, the outcome was tested related to time since the admission until death/end of the study period. For people who did not die, end of July 2020 was included as the final day of research; therefore, the study ended. The Kaplan–Meier survival curves were performed and differences in survival rates were analyzed by log-rank test. Univariate Cox regression was conducted to detect the hazard ratio (HR) of different comorbidities and variables with statistically different HR were included in a multivariate Cox regression model and adjusted for age and sex. In all statistical tests p-value <0.05 was considered statistically significant.

## Results

Screening of 447 patients’ records was conducted and 8 cases were excluded due to their age (< 18 years old) then analysis of 439 cases was performed. Patients with comorbidities represented 61.7% of all cases as demonstrated in ***figure 1***.

**Figure (1):**
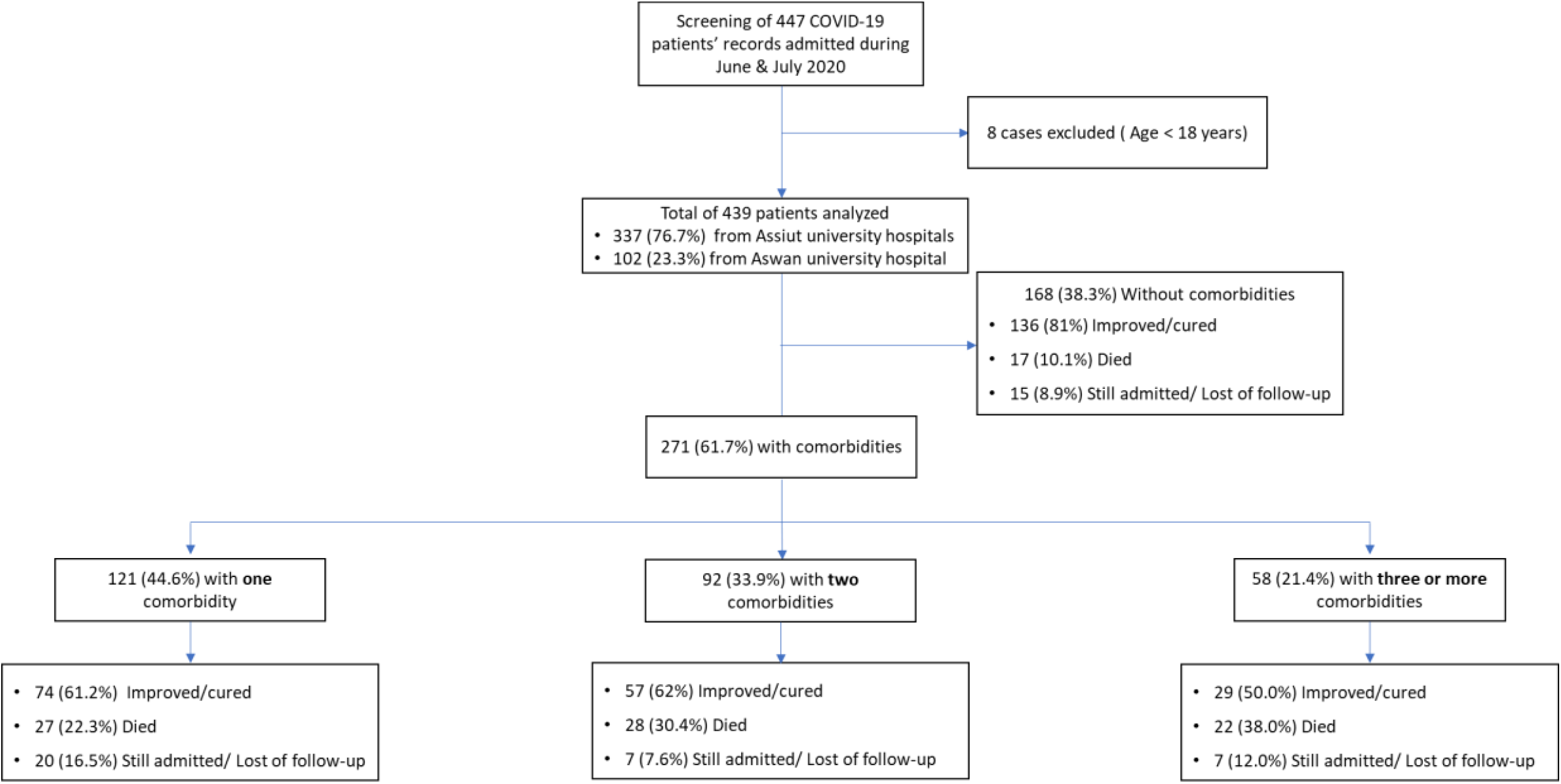
Flowchart of COVID-19 patients included in the study stratified according to number of comorbidities.

The mean age of studied patients was 51.2 ± 17.2 years old and it was higher in patients with comorbidities (p < 0.001). Fever, and lower respiratory tract (LRT) symptoms especially dry cough were the most frequent symptoms (74.3% and 74.7% respectively). GIT symptoms were present in 21.2% of patients, while neurological symptoms in 43.8%. When comparing symptoms of two groups, constitutional symptoms especially myalgia and LRT symptoms such as dyspnea were significantly higher in patients with comorbidities (P < 0.05). CT-chest showed bilateral GGO in most of cases (82.6%). ***(Table 1)***

**Table 1.** **Demographic and clinical characteristics of COVID-19 patients included in the study (n=439)**

|  | All patients<br>(n=439) | Patients without<br>comorbidity<br>(n=168) | Patients with<br>comorbidities<br>(n=271) | p-value* |
| --- | --- | --- | --- | --- |
| Age (years) |  |  |  |  |
| 18 – 39 | 122 (27.8%) | 96 (57.1%) | 26 (9.6%) | < 0.001^ |
| 40 – 59 | 147 (33.5%) | 51 (30.4%) | 96 (35.4%) |  |
| 60 – 79 | 153 (34.9%) | 19 (11.3%) | 134 (49.4%) |  |
| ≥ 80 | 17 (3.9%) | 2 (1.2%) | 15 (5.5%) |  |
| Mean ± SD | 51.2 ± 17.2 | 39.4 ± 15.0 | 58.5 ± 14.0 | < 0.001^ |
| Gender, n(%) |  |  |  |  |
| Male | 224 (51%) | 86 (51.2%) | 138 (50.9%) | 0.956 |
| Female | 215 (49%) | 82 (48.8%) | 133 (49.1%) |  |
| Presenting symptoms, n (%) |  |  |  |  |
| Fever | 326 (74.3%) | 118 (70.2%) | 208 (76.8%) | 0.129 |
| Constitutional symptoms,<br>n(%) | 177 (40.3%) | 55 (32.7%) | 122 (45.0%) | 0.011 ^ |
| Fatigue | 125 (28.5%) | 42 (25%) | 83 (30.6%) | 0.204 |
| Bone pain | 18 (4.1%) | 7 (4.2%) | 11 (4.1%) | 0.956 |
| Myalgia | 63 (14.4%) | 17 (10.1%) | 46 (17%) | 0.045 ^ |
| Anorexia | 48 (10.9%) | 12 (7.1%) | 36 (13.3%) | 0.051 |
| URT symptoms, n (%) | 110 (25.1%) | 37 (22%) | 73 (26.9%) | 0.248 |
| Sore throat | 96 (21.9%) | 31 (18.5%) | 65 (24%) | 0.173 |
| Nasal congestion | 24 (5.5%) | 11 (6.5%) | 13 (4.8%) | 0.433 |
| LRT symptoms, n (%) | 328 (74.7%) | 113 (67.3%) | 215 (79.3%) | 0.005 ^ |
| Cough | 293 (66.7%) | 105 (62.5%) | 188 (69.4%) | 0.137 |
| Sputum | 116 (26.4%) | 24 (14.3%) | 92 (33.9%) | < 0.001 ^ |
| ***Dyspnea*** | 208 (47.4%) | 51 (30.4%) | 157 (57.9%) | <0.001 ^ |
| **GIT symptoms, n(%)** | 93 (21.2%) | 36 (21.4%) | 57 (21%) | 0.922 |
| ***Abdominal pain*** | 11 (3.5%) | 6 (3.6%) | 5 (1.8%) | 0.261 |
| ***Nausea/Vomiting*** | 24 (5.5%) | 6 (3.6%) | 18 (6.6%) | 0.169 |
| ***Diarrhea*** | 69 (15.7%) | 31 (18.5%) | 38 (14%) | 0.215 |
| **neurological symptoms, n (%)** | 192 (43.8%) | 74 (44%) | 118 (43.5%) | 0.917 |
| ***Headache*** | 83 (18.9%) | 27 (16.1%) | 56 (20.7%) | 0.232 |
| ***Dizziness*** | 63 (14.4%) | 27 (16.1%) | 36 (13.3%) | 0.418 |
| ***Vertigo*** | 3 (0.7%) | 1 (0.6%) | 2 (0.7%) | 0.860 |
| ***Loss of smell*** | 35 (8%) | 15 (8.9%) | 20 (7.4%) | 0.560 |
| ***Loss of taste*** | 25 (5.7%) | 10 (6%) | 15 (5.5%) | 0.854 |
| **CT chest findings, n (%)** |  |  |  |  |
| **Normal** | 16 (4.9%) | 10 (9.2%) | 6 (2.8%) | 0.124 |
| **Bilateral GGO** | 270 (82.6%) | 87 (79.8%) | 183 (83.9%) |  |
| **Unilateral GGO** | 5 (1.5%) | 2 (1.8%) | 3 (1.4%) |  |
| **GGO and consolidations** | 36 (11%) | 10 (9.2%) | 26 (12%) |  |

**URT= upper respiratory tract; LRT=lower respiratory tract; GIT=gastrointestinal tract; CT= computed tomography; GGO=ground glass opacities**
**\* Students' T-test and Chi-square test were used.**
**^ significant p-value.**

The most frequent comorbidities in studied group were cardiovascular diseases (69%) of patients with comorbidities followed by DM (54.2%). ***(Table 2)***

**Table 2.** **Frequency (number and percent) of different comorbidities among 439 COVID-19 patients**

| Comorbidity | Number | % from cases with comorbidities | % from all cases |
| --- | --- | --- | --- |
| **Diabetes Miletus** | **147** | **54.2** | **33.5** |
| **Cardiovascular Diseases** | **187** | **69.0** | **42.6** |
| Hypertension | 131 (70.0%) | 63.4 | 39.2 |
| Ischemic heart disease | 8 (4.3%) | 3.0 | 1.8 |
| Hypertension & IHD | 41 (21.9%) | 15.1 | 9.3 |
| AF | 4 (2.1%) | 1.5 | 0.91 |
| Valvular disease | 2 (1.1 %) | 0.7 | 0.46 |
| Congenital cyanotic heart disease | 1 (0.53%) | 0.37 | 0.23 |
| **Chronic pulmonary diseases** | **31** | **11.4** | **7.1** |
| COPD | 12 (38.7%) | 4.4 | 2.7 |
| Asthma | 10 (32.3%) | 3.7 | 2.2 |
| ILD | 3 (9.7%) | 1.1 | 0.68 |
| SRBD | 3 (9.7%) | 1.1 | 0.68 |
| Previous pulmonary embolism | 2 (6.5%) | 0.74 | 0.46 |
| Pleural effusion | 1 (3.2%) | 0.37 | 0.23 |
| **Neuropsychiatric diseases** | **31** | **11.4** | **7.1** |
| Old CVS | 16 (51.6%) | 5.9 | 3.6 |
| Brain tumors | 4 (12.9%) | 1.5 | 0.91 |
| Epilepsy | 3 (9.7%) | 1.1 | 0.68 |
| Parkinson's disease dementia (PDD) | 2 (6.5%) | 0.74 | 0.46 |
| MS | 2 (6.5%) | 0.74 | 0.46 |
| Parkinson's disease (PD) | 1 (3.2%) | 0.37 | 0.23 |
| Alzheimer's Dementia | 1 (3.2%) | 0.37 | 0.23 |
| MG | 1 (3.2%) | 0.37 | 0.23 |
| Brain abscess | 1 (3.2%) | 0.37 | 0.23 |
| **Kidney disease** | **21** | **7.7** | **4.8** |
| Chronic kidney disease | 18 (85.7%) | 6.6 | 4.1 |
| ESRF on RD | 2 (9.5%) | 0.74 | 0.46 |
| Renal transplant | 1 (4.8%) | 0.37 | 0.23 |
| **Liver Disease** | **15** | **5.5** | **3.4** |
| HCV | 7 (46.7%) | 2.6 | 1.6 |
| HCV with Cirrhosis | 5 (33.3%) | 1.8 | 1.1 |
| Portal hypertension with bleeding varices | 1 (6.7%) | 0.37 | 0.23 |
| Autoimmune hepatitis | 1 (6.7%) | 0.37 | 0.23 |
| Liver transplant | 1 (6.7%) | 0.37 | 0.23 |
| **Metabolic and Endocrinal Diseases** | **10** | **3.7** | **2.3** |
| Hypothyroidism | 8 (80%) | 3.0 | 1.8 |
| Gout | 2 (20%) | 0.74 | 0.46 |
| **Autoimmune Diseases** | **7** | **2.6** | **1.6** |
| SLE | 3 (42.9%) | 1.1 | 0.68 |
| RA | 1 (14.3%) | 0.37 | 0.23 |
| Autoimmune vasculitis | 1 (14.3%) | 0.37 | 0.23 |
| Behcet's disease | 1 (14.3%) | 0.37 | 0.23 |
| Scleroderma | 1 (14.3%) | 0.37 | 0.23 |
| **Other conditions/risk factors** | **9** | **3.3** | **2.1** |
| Malignancy <sup>a</sup> | 3 (18.8%) | 1.1 | 0.68 |
| Chronic bilateral LL ischemia | 3 (18.8%) | 1.1 | 0.68 |
| Pregnancy | 3 (18.8%) | 1.1 | 0.68 |

<sup>a</sup> Acute Leukemia, Lymphoma & breast cancer on chemotherapy.
IHD: ischemic heart disease. AF: atrial fibrillation. COPD: chronic obstructive pulmonary disease. ILD: interstitial lung diseases. SRBD: sleep related breathing disorders. ESRF on RD: end stage renal failure on regular dialysis. CVS: cerebrovascular stroke. HCV= hepatitis C virus; HCV= hepatitis C virus;; CVS= cerebrovascular stroke; MS= multiple sclerosis; MG= myasthenia gravis; SLE= systemic lupus erythematosus; RA= rheumatoid arthritis

Patients with comorbidities had significantly worse laboratory parameters. Detailed data were included in ***Table 3***.

**Table 3.** **Laboratory data of COVID-19 patients included in the study**

|  | Cases<br>with<br>available<br>data | All patients | Patients without<br>comorbidity | Patients with<br>comorbidities | p-value* |
| --- | --- | --- | --- | --- | --- |
| CBC |  |  |  |  |  |
| Hemoglobin (g/dl) | 393 | 12.2 ± 2.0 | 12.6 ± 1.9 | 11.9 ± 2.0 | 0.001 ^ |
| Anemic (HB < 12) |  | 178 (45.3%) | 54 (35.8%) | 124 (51.2%) | 0.003 ^ |
| WBCs (10<sup>3</sup>/ul) | 405 | 9.2 ± 5.5 | 8.3 ± 4.8 | 9.8 ± 5.9 | 0.003 ^ |
| Low (<4) |  | 46 (11.4%) | 23 (14.8%) | 21 (9.2%) | 0.014 ^ |
| High (>10) |  | 143 (35.3%) | 42 (27.1%) | 101 (40.4%) |  |
| Neutrophil (%) | 315 | 68.1 ± 20.4 | 63.5 ± 20.9 | 70.5 ± 19.7 | 0.013 ^ |
| Low (< 40) |  | 26 (8.3%) | 13 (11.8%) | 13 (6.3%) | 0.029 ^ |
| High (>75) |  | 144 (45.7%) | 40 (36.4%) | 104 (50.7%) |  |
| Lymphocytes (%) | 407 | 19.9 ± 14.6 | 23.0 ± 15.6 | 18.1 ± 13.7 | 0.001 ^ |
| Low (< 20) |  | 242 (59.8%) | 82 (53.6%) | 160 (63.5%) | 0.085 |
| Lymphocytes (10<sup>3</sup>/ul) | 407 | 1.4 ± 0.98 | 1.5 ± 0.99 | 1.4 ± 0.98 | 0.301 |
| Low (< 1.5) |  | 256 (62.9%) | 92 (59.7%) | 164 (64.8%) | 0.542 |
| Platelets (10<sup>3</sup>/ul) | 391 | 275.2 ± 139.1 | 283.9 ± 118.7 | 269.8 ± 150.5 | 0.177 |
| Low (< 140) |  | 37 (9.5%) | 4 (2.7%) | 33 (13.7%) | < 0.001^ |
| High (> 450) |  | 37 (9.5%) | 10 (6.7%) | 27 (11.2%) |  |
| INR | 256 | 1.1 ± 0.32 | 1.0 ± 0.12 | 1.1 ± 0.39 | 0.092 |
| Ferritin (ng/ml) | 262 | 749.0 ± 997.4 | 698.9 ± 1137.5 | 777.9 ± 909.0 | 0.002 ^ |
| High (>291) |  | 165 (63%) | 50 (52.1%) | 115 (69.3%) | 0.005 ^ |
| D-dimer (mg/L) | 297 | 2.5 ± 8.7 | 1.3 ± 3.4 | 3.5 ± 11.1 | < 0.001 ^ |
| High (> 0.55) |  | 180 (60.6%) | 53 (41.7%) | 127 (74.7%) | <0.001 ^ |
| CRP (mg/dl) | 289 | 57.8 ± 74.1 | 43.0 ± 74.5 | 68.3 ± 72.2 | < 0.001 ^ |
| High (>1) |  | 282 (97.6%) | 114 (95%) | 168 (99.4%) | 0.016 ^ |
| PCR |  |  |  |  |  |
| Positive | 416 | 354 (85.1%) | 144 (88.9%) | 210 (82.7%) | 0.083 |
| Negative |  | 43 (10.6%) | 18 (11.1%) | 44 (17.3%) |  |

CBC= complete blood picture; WBCs= white blood cells; INR= international normalized ratio; PCR= Polymerase chain reaction.
\* Students' T-test. Mann-Whitney and Chi-square test were used.
^ significant p-value.

ICU admission was higher in patients with comorbidities as 35.8% of them need ICU admission compared to only 16.4% of those without any comorbidity (P<0.001). Among different comorbidities 45.4% of CVD cases were admitted in ICU followed by DM cases (40.8%) ***(Figure 2)***. Also, patients with comorbidities needed invasive mechanical ventilation more than those without comorbidity (31 vs. 10.7%, P<0.001). ***(Table 4)***

**Table 4.** **Clinical outcomes of COVID-19 patients included in the study (n=439)**

|  | All patients<br>(n=439) | Patients without<br>comorbidity<br>(n=168) | Patients with<br>comorbidities<br>(n=271) | p-value* |
| --- | --- | --- | --- | --- |
| **ICU Admission** | 124 (34.1%) | 28 (16.4%) | 96 (35.8%) | < 0.001 ^ |
| **Oxygen therapy** |  |  |  |  |
| *No need* | 130 (29.6%) | 76 (45.2%) | 54 (19.9%) | < 0.001 ^ |
| *Simple face mask* | 134 (30.5%) | 53 (31.5%) | 81 (29.9%) |  |
| *Venturi mask* | 45 (10.3%) | 14 (8.3%) | 31 (11.4%) |  |
| *Non rebreathing mask* | 11 (2.5%) | 3 (1.8%) | 8 (3.0%) |  |
| *Noninvasive MV* | 17 (3.9%) | 4 (2.4%) | 13 (4.8%) |  |
| *Invasive MV* | 102 (23.2%) | 18 (10.7%) | 84 (31.0%) |  |
| **Outcome <sup>a</sup>** |  |  |  |  |
| *Cured/Improved* | 296 (67.4%) | 136 (81%) | 160 (59%) | < 0.001 ^ |
| *Died* | 94 (21.4%) | 17 (10.1%) | 77 (28.4%) |  |
| **Duration of hospital stay** | 8.4 ± 6.1 | 8.5 ± 6.1 | 8.4 ± 6.0 | 0.853 |

ICU= intensive care unit
\* Students' T-test and Chi-square test were used.
^ significant p-value.
<sup>a</sup> 49 patients (11.2%) were still admitted until the end of study period or lost follow-up.

**Figure (2):**
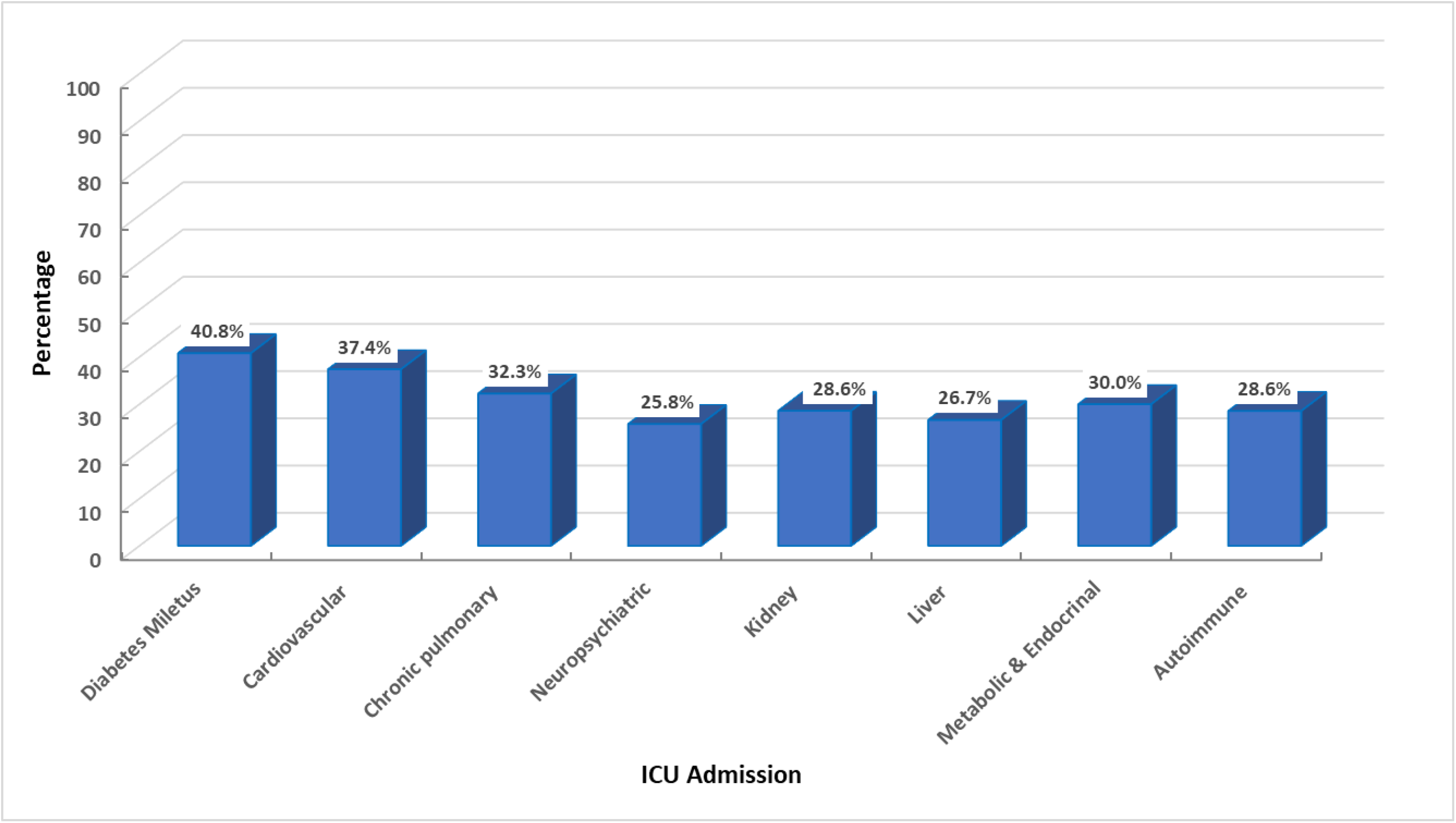
ICU admission according to comorbidities.

Recovery was recorded in nearly 68% of admitted COVID-19 cases (improved and discharged from hospital) with significant lower frequency of cure in patients with comorbidities (59% vs. 81%, P<0.001) and death rate was significantly higher in cases with comorbidities (P< 0.001) ***(Table 4)***. Death rate in different comorbidity groups was illustrated in ***figure 3***.

**Figure (3):**
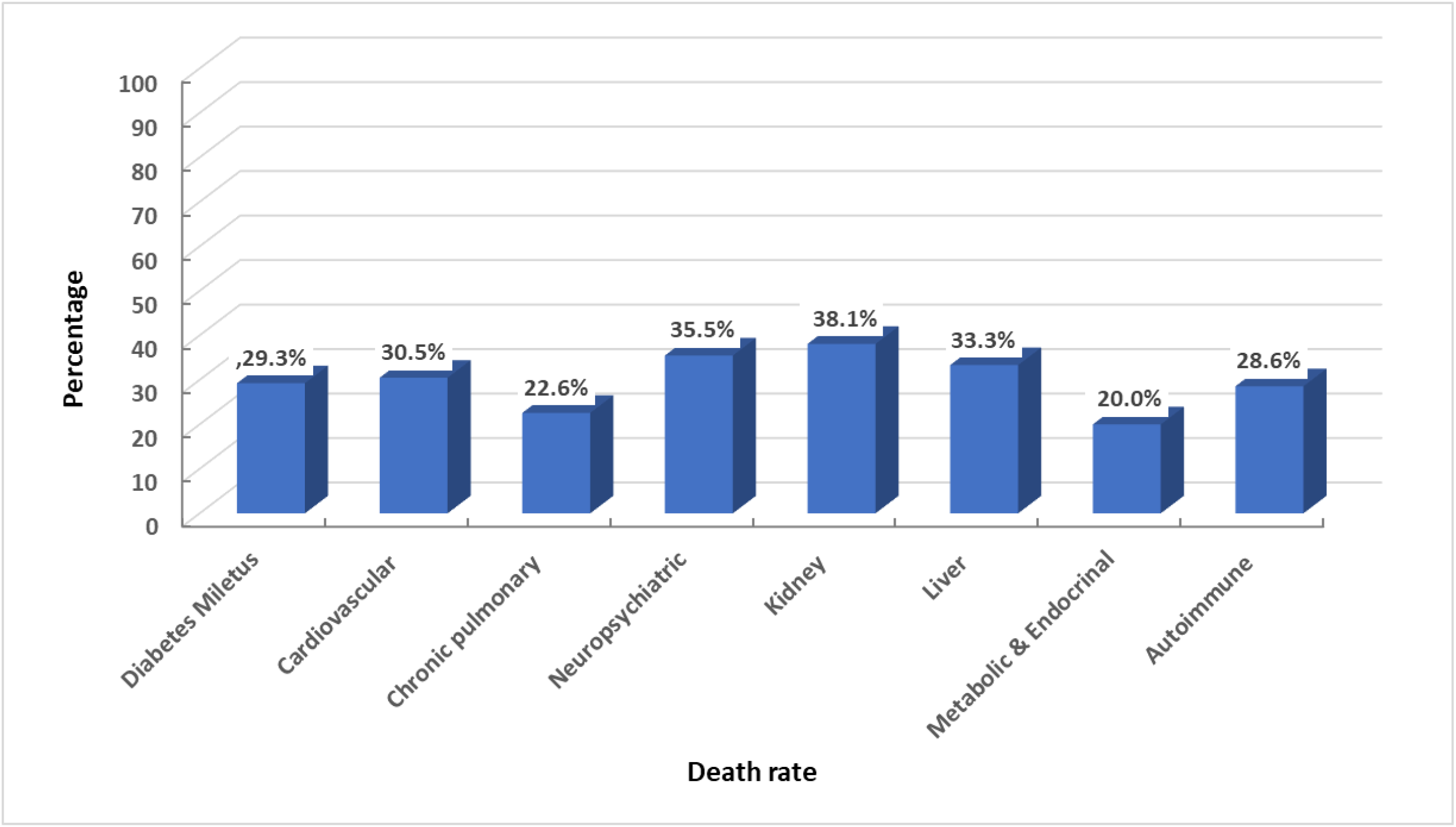
Death rate according to comorbidities.

Kaplan-Maier survival curves showed significant difference in survival rates in cases with pre-existing CVD and neurological diseases in which the survival rates are lower than those without disease (P<0.002 and 0.001 respectively). Also, patients without any comorbidity showed higher survival rate than those with one, two or three or more comorbidities (74.2%, 34.5%, 34.3% and 35.5% respectively). ***(Figures 4&5)***

**Figure (4):**
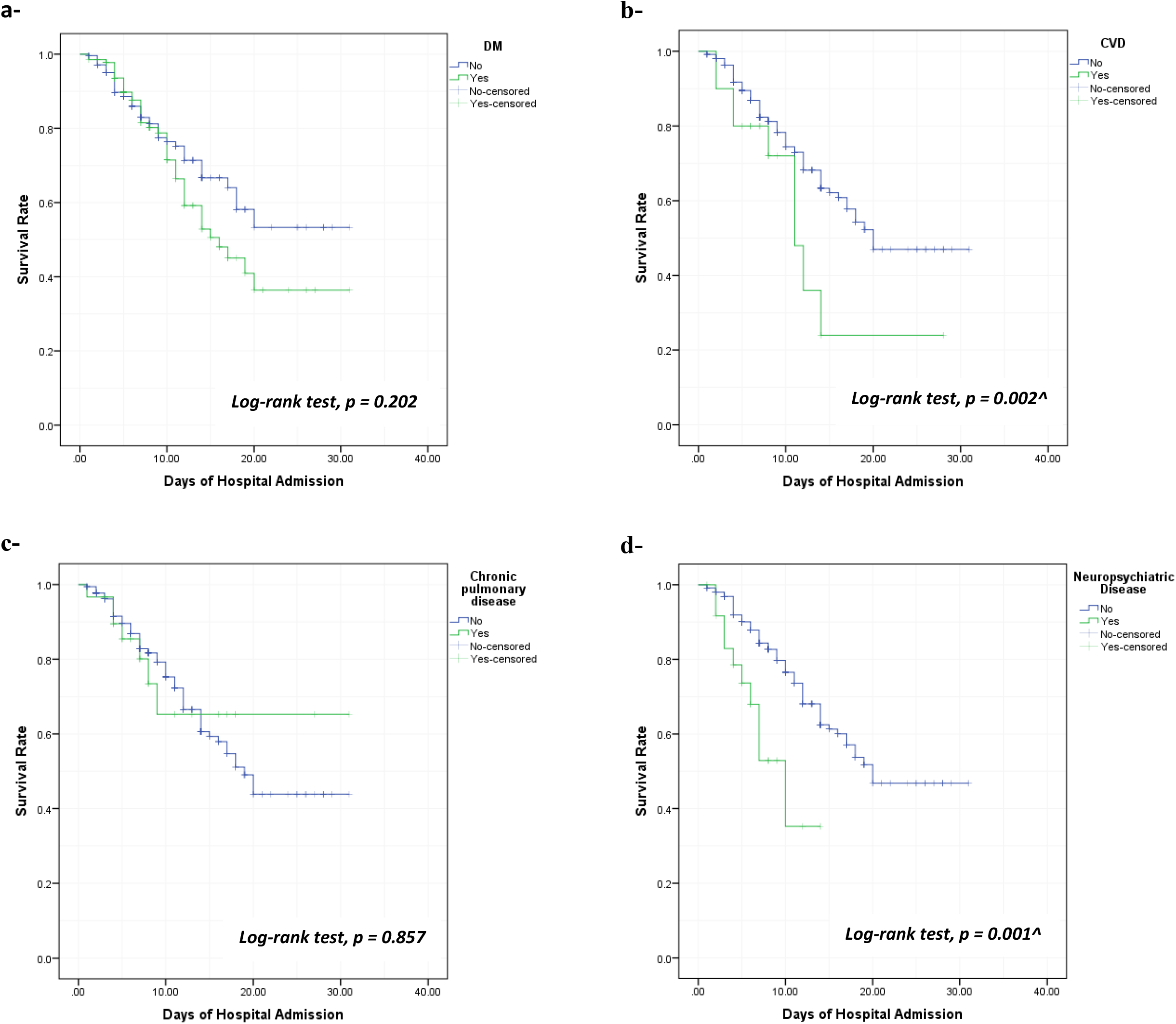
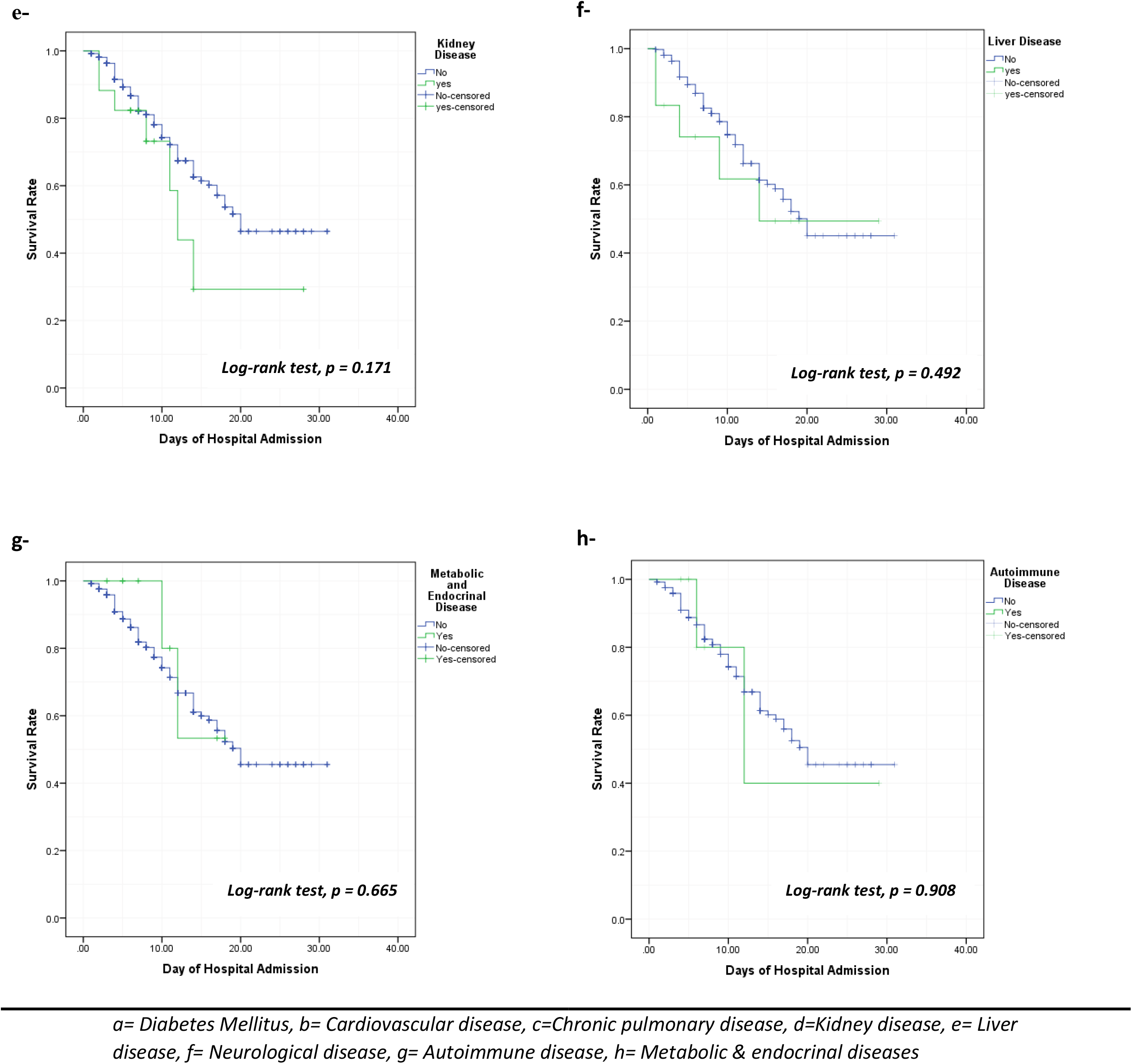
Kaplan-Meir survival curve of COVID-19 cases based on different associated comorbidities (n=439).

**Figure (5):**
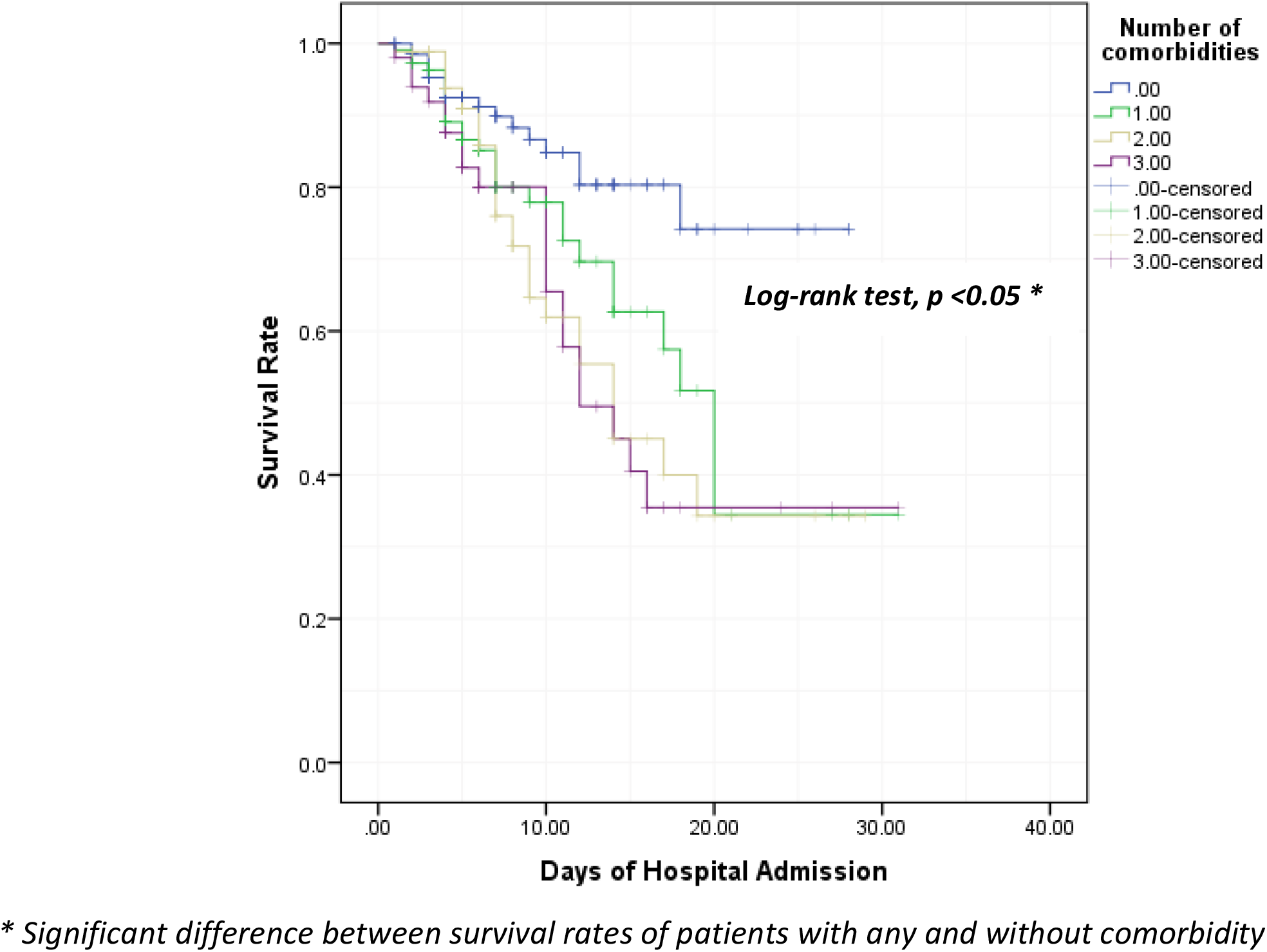
Kaplan-Meir survival curve of COVID-19 cases included in the study stratified according to number of comorbidities (n=439).

In univariate cox regression model, the older ages had higher risk of death than lower age groups as HR for 60 – 79 age group was 3.9 (95% CI. 2 – 7.8) and in cases > 80 years was 10.1 (95% CI. 4-25.6). For different comorbidities, cases with CVDs and neurological diseases showed significant higher risk of death than others as HR in CVDs was 1.9 (95% CI. 1.2 – 1.9) and in neurological diseases was 2.9 (95% CI. 1.5 – 5.6). Also, number of comorbidities showed significant higher risk than those without any comorbidity as those with one comorbidity has a HR of 2 (95% CI. 1.1 – 3.7) and with two comorbidities the HR as 2.6 (95% CI. 1.4 – 4.7) while those with 3 or more comorbidities the HR as 2.9 (95% CI. 1.5 – 5.6). After adjustment of significant factors in multivariate cox regression, only higher age groups and neurological diseases showed higher risk for death. ***(Table 5)***

**Table 5.**
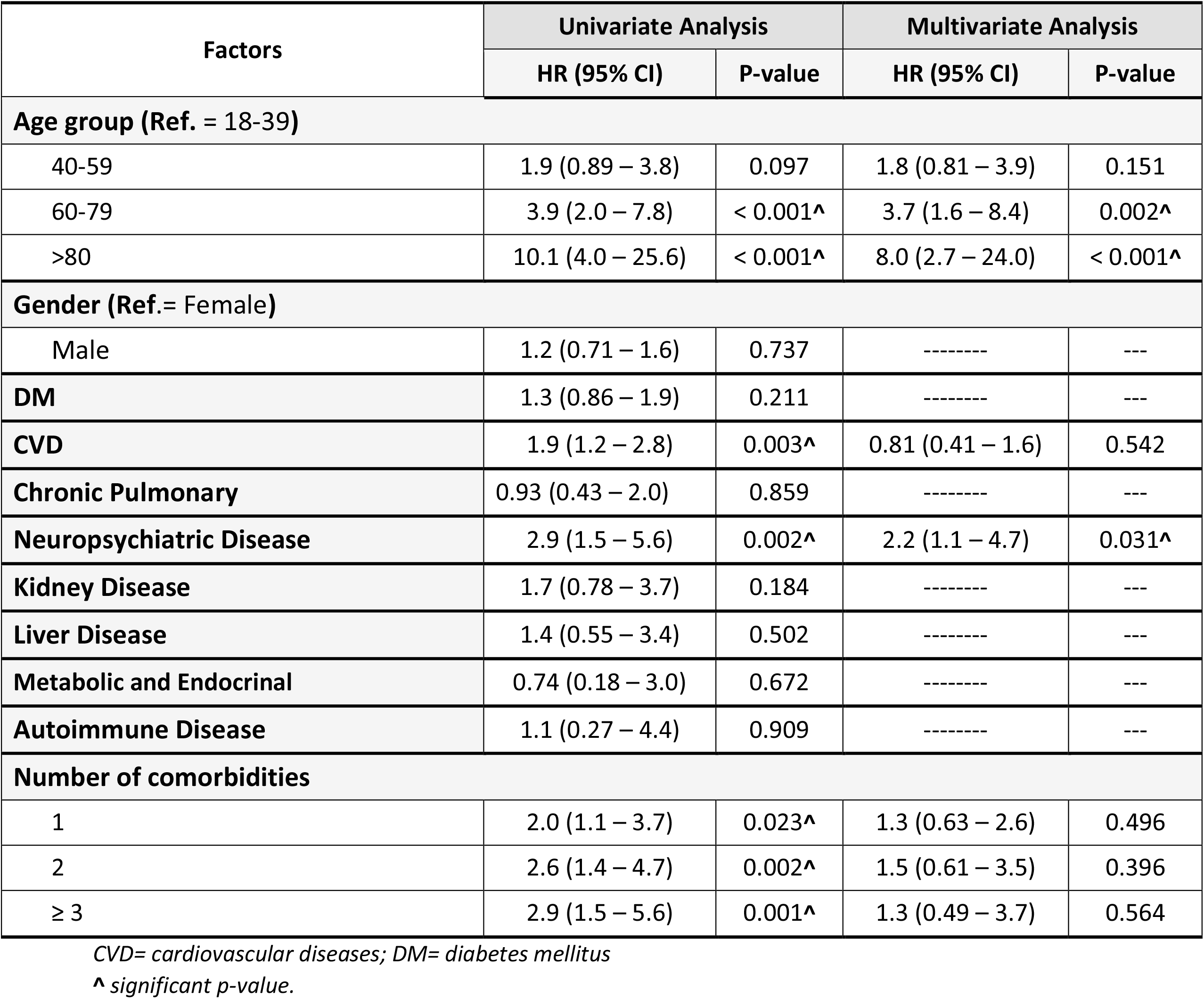
**Cox regression analysis of factors associated with overall survival in included COVID-19 cases in the study (n=439)**

| Factors | Univariate Analysis |  | Multivariate Analysis |  |
| --- | --- | --- | --- | --- |
|  | HR (95% CI) | P-value | HR (95% CI) | P-value |
| **Age group (Ref. = 18-39)** |  |  |  |  |
| 40-59 | 1.9 (0.89 – 3.8) | 0.097 | 1.8 (0.81 – 3.9) | 0.151 |
| 60-79 | 3.9 (2.0 – 7.8) | < 0.001^ | 3.7 (1.6 – 8.4) | 0.002^ |
| >80 | 10.1 (4.0 – 25.6) | < 0.001^ | 8.0 (2.7 – 24.0) | < 0.001^ |
| **Gender (Ref.= Female)** |  |  |  |  |
| Male | 1.2 (0.71 – 1.6) | 0.737 | ----- | --- |
| **DM** | 1.3 (0.86 – 1.9) | 0.211 | ----- | --- |
| **CVD** | 1.9 (1.2 – 2.8) | 0.003^ | 0.81 (0.41 – 1.6) | 0.542 |
| **Chronic Pulmonary** | 0.93 (0.43 – 2.0) | 0.859 | ----- | --- |
| **Neuropsychiatric Disease** | 2.9 (1.5 – 5.6) | 0.002^ | 2.2 (1.1 – 4.7) | 0.031^ |
| **Kidney Disease** | 1.7 (0.78 – 3.7) | 0.184 | ----- | --- |
| **Liver Disease** | 1.4 (0.55 – 3.4) | 0.502 | ----- | --- |
| **Metabolic and Endocrinal** | 0.74 (0.18 – 3.0) | 0.672 | ----- | --- |
| **Autoimmune Disease** | 1.1 (0.27 – 4.4) | 0.909 | ----- | --- |
| **Number of comorbidities** |  |  |  |  |
| 1 | 2.0 (1.1 – 3.7) | 0.023^ | 1.3 (0.63 – 2.6) | 0.496 |
| 2 | 2.6 (1.4 – 4.7) | 0.002^ | 1.5 (0.61 – 3.5) | 0.396 |
| ≥ 3 | 2.9 (1.5 – 5.6) | 0.001^ | 1.3 (0.49 – 3.7) | 0.564 |

CVD= cardiovascular diseases; DM= diabetes mellitus
^ significant p-value.

## Discussion

Multiple comorbidities are associated with COVID-19 disease and may contribute to its progression or poor outcome. The current study included 439 moderate to severe COVID-19 patients of them 61.7% had one or more comorbid diseases. The mean age of the studied population was 51.2 years with no gender predilection. Of them, 34.1% needed ICU admission and 21.4% were deceased with mean hospital stay 8.4 days.

In this study, as previously reported in reviews and metanalysis(5, 8, 9) cardiovascular diseases, including hypertension,(42.6%) and diabetes mellitus(33.5%) were the most common comorbidities associated with COVID-19 infection followed by chronic chest diseases and neuropsychiatric disorders (7.1%). This may be attributed to: A) The older age of patients with comorbid diseases. Based on the current information, the elderly, a vulnerable population, with chronic health conditions such as diabetes and cardiovascular or lung disease not only a higher risk of COVID-19 infection(10) but also an increased risk of getting severe illness or even death if they become infected(11). The weaknesses of advanced age are related to the function defense cells T and B, and to the excess production of type 2 cytokines, which can lead to a prolonged pro-inflammatory response, leading to unfortunate results(12). B) Common pathogenesis as chronic disorders collaborate several baseline topographies with communicable diseases, for instance the pro-inflammatory status, and the attenuation of the innate immune response. For instance, diabetes occurs in part because the accumulation of triggered innate immune cells leads to the release of inflammatory markers, principally IL-1β and Tumor necrosis factor α, that promote generalized resistance to insulin and destruction of β-cell (13). C) Immunity depletion by impairing macrophage and lymphocyte function which may make individuals more susceptible to infectious diseases and its complications(3). D) Up-regulation of angiotensin-converting enzyme 2 (ACE2) genes expression in different parts of the body, such as heart and lungs, in patients with diabetes, or CVD, increasing the susceptibility to SARS-CoV-2 infection and the risk of disease aggravation as it has been identified as an important functional receptor for SARS-CoV-2 invasion(4, 14).

In the current study, cases with severe disease requiring ICU admission (35.8% compared to 16.4%), increased need for oxygen therapy, ventilatory support, and almost triple times death rate were significantly associated with patients recording comorbidities. However, by using univariate cox regression model, only cases with CVDs (HR 1.9; 95% CI 1.2 – 2.8; P<0.003) and neurological diseases (HR 2.9; 95% CI 1.5 – 5.6; P<0.002) showed significant higher risk of death than others. After adjustment of significant factors in multivariate cox regression, neurological diseases showed higher risk for death.

Cardiovascular comorbid conditions are associated with a variety of the worse outcomes for COVID-19 in literatures (15-17). Moreover, association between hypertension and co-existing other comorbidities such as HIV, kidney, or other cardiovascular diseases raises the hazard rate more than the other comorbidities(17). In this study, 37.4% of patients with CVDs including hypertension admitted to ICU and 30.5% of them were deceased. M. Li et al. found that the levels of inflammation indicators like CRP, serum ferritin and ESR were increased in COVID-19 patients and associated with the severity of the disease, furthermore, the levels of these indicators in the patients with CVD were ominously higher than those without CVD which indicate that COVID-19 cases with CVD had additional potential to practice an inflammatory storm, which ultimately clues to prompt worsening of these patients’ conditions. Through the course of contagion, inflammation of the lung tissue inhibits the exchange of oxygen in the alveoli, progressing to generalized tissue hypoxia, which triggers the fibrinolytic system. Moreover, compared with the non-CVD group, the ranks of D-dimer and FIB were greater among CVD group, which indicates that they were more susceptible to hyper-coagulability. A hyper-coagulable state raises the hazard of pulmonary embolism, which can explain the sudden occurrence of complications such as hypoxia and heart failure(16). All those results were harmonized with the current study which displayed significantly higher levels of all inflammatory markers as well as D-dimer level in patients with comorbidities, however, due to the overlap between different conditions (33.9% of patients had 2 comorbid conditions and 21.4% had three or more), we couldn’t evaluate patients with CVD alone.

Unlike most of the published literatures, even a recent observational study carried out in the same geographical area which stated that “pre-existing DM may predict unfortunate 30 days-in hospital outcome” (18), diabetes mellitus was not found to be a risk factor for death in patients with COVID-19 (HR 1.3; 95% CI0.86 – 1.9; p< 0.211) despite that 40.8% of patient with DM were admitted to ICU meanwhile, 29.3% were deceased. In a cohort study of 7337patients with COVID-19, it was shown that those with type 2 diabetes not only prerequisite augmented interferences for their stay in hospital versus those that were non-diabetic, but also had an increased mortality rate(19). In the current study, in contrast with the above-mentioned ones, we considered DM as a comorbidity when it is self-reported by patient (previous diagnosis and regular treatment) not on the base line random blood glucose at admission. According to Yan Y. et al. non-survivors with diabetes had higher levels of leukocyte count, neutrophil count, C-reactive protein, pro-calcitonin, ferritin, receptors of interleukin-2, interleukin-6, TNF-α, and lower lymphocytic count than survivors, which indicated diabetic non-survivors had more sever inflammatory response(20).

Despite the notable COVID-19 cases presentation with acute cerebrovascular accidents(23), pre-existing neurological conditions with their wide spectrum of diseases also related to COVID-19 infection and mortality. One survival analysis in 2070 Brazilian patients found that a risk of 3.9 times in people with neurologic disease (95% CI 1.9–7.8; P<0.001)(4). The increased severity of COVID-19 among cases with cerebrovascular illness may be attributed to coexistence of cerebrovascular diseases with other risk factors such as older age, cardiovascular diseases, and diabetes mellitus. Besides, the existence of brain medullary cardiorespiratory or autonomic nervous system dysfunction precipitate to blood pressure fluctuation and dysfunction in the respiratory system increasing the hazard of acquiring opportunistic infections. Moreover, the relative immobility in post-stroke patients, increases the risk for hyper-coagulable state(21).

Among other comorbidities, chronic pulmonary diseases including obstructive pulmonary disease (COPD) and moderate to severe bronchial asthma are at higher risk of sever COVID-19 progression since this virus affects their respiratory tracts, leading to increased bronchospasm, pneumonia, and acute respiratory distress(10). However, in contrast to the above-mentioned data, another study found that asthma was a protective factor. Asthma may not be a risk factor because of reduced angiotensin-converting enzyme-2 (ACE2) gene expression in airway cells of asthma patients that would be expected to decrease the severity of SARS-CoV-2 infection which uses ACE2 as its cellular receptor(22). We suggest that the degree of asthma control, medications used for asthma control, difference in patient ages and the coexisting conditions may explain this disagreement. Besides, a high-quality cohort study with longer time frame and larger number of patients is needed to explain asthma COVID-19 interaction.

Unfortunately, a smaller number of patients with other comorbidities such as chronic kidney, liver, metabolic, endocrine, and autoimmune diseases as well as patients with malignancy were included in this study. However, the highest death rate among all the comorbid conditions in this study was (38.1%) of patients with kidney diseases. Cases with liver disorders also had relatively high mortality rate (33.3%). In literatures, the rate of mortality in COVID-19 patients with CKD and Liver diseases was found to be (53.33%) and (17,65%) respectively (23). Chronic kidney diseases are associated with dysregulated inflammation, immune system, and levels of ACE2 receptors in kidneys which may explain the severity and mortality due to COVID-19 in patients with CKD(23). Worth notice that, in this study, patients with comorbidities had significantly higher renal function tests together with elevated inflammatory biomarkers.

Limitations of the study:

- The retrospective design of the study with the heterogeneity of data recoding in different hospital sectors, b) the changing treatment protocol and admission policy during the study period, c) some subjects having more than one underlying comorbidity, and d) by the end of study some included patients are still admitted with unclear outcome yet.

## Conclusion

Coexistence of cardiovascular comorbid conditions including hypertension or neurological diseases together with COVID-19 infections carries higher risks of mortality. However, other comorbidities such as diabetes mellitus, chronic pulmonary or kidney diseases may also contribute to increased COVID-19 severity. Special attention should be taken during dealing with such cases. Future studies with larger number of patients in wider varieties of comorbidities and longer duration evaluating their impact on survival are desired.

## Data Availability

The data referred to in the manuscript are available.

## References

1. Coronavirus disease (COVID-19) outbreak: World Health Organization; 2020 [cited 2020 September 4]. Available from: https://covid19.who.int/.

2. Ghoniem A, Abdellateef A, Osman AI, Elsayed HH, Elkhayat H, Adel W. A tentative guide for thoracic surgeons during COVID-19 pandemic. The Cardiothoracic Surgeon. 2020;28(1):16.

3. Badawi A, Ryoo SG. Prevalence of comorbidities in the Middle East respiratory syndrome coronavirus (MERS-CoV): a systematic review and meta-analysis. Int J Infect Dis. 2016;49:129-33.

4. Sousa GJB, Garces TS, Cestari VRF, Florencio RS, Moreira TMM, Pereira MLD. Mortality and survival of COVID-19. Epidemiology and infection. 2020;148:e123.

5. Yang J, Zheng Y, Gou X, Pu K, Chen Z, Guo Q, et al. Prevalence of comorbidities and its effects in patients infected with SARS-CoV-2: a systematic review and meta-analysis. Int J Infect Dis. 2020;94:91-5.

6. Simonnet A, Chetboun M, Poissy J, Raverdy V, Noulette J, Duhamel A, et al. High Prevalence of Obesity in Severe Acute Respiratory Syndrome Coronavirus-2 (SARS-CoV-2) Requiring Invasive Mechanical Ventilation. Obesity (Silver Spring). 2020;28(7):1195-9.

7. Bajgain KT, Badal S, Bajgain BB, Santana MJ. Prevalence of comorbidities among individuals with COVID-19: A rapid review of current literature. American Journal of Infection Control. 2020.

8. Zhou Y, Yang Q, Chi J, Dong B, Lv W, Shen L, et al. Comorbidities and the risk of severe or fatal outcomes associated with coronavirus disease 2019: A systematic review and meta-analysis. Int J Infect Dis. 2020;99:47-56.

9. Sanyaolu A, Okorie C, Marinkovic A, Patidar R, Younis K, Desai P, et al. Comorbidity and its Impact on Patients with COVID-19. SN comprehensive clinical medicine. 2020:1-8.

10. CDC. Coronavirus (COVID-19): symptoms of coronavirus: Centers for Disease Control and Prevention; 2020 [cited 2020 April 18]. Available from: https://www.cdc.gov/coronavirus/2019-ncov/symptoms-testing/symptoms.html.

11. BCCDC. COVID-19 vulnerable populations.: British Columbia Centre for Disease Control; 2020 [cited 2020 April 18]. Available from: http://www.bccdc.ca/health-info/diseases-conditions/covid-19/vulnerable-populations.

12. Opal SM, Girard TD, Ely EW. The immunopathogenesis of sepsis in elderly patients. Clinical infectious diseases : an official publication of the Infectious Diseases Society of America. 2005;41 Suppl 7:S504-12.

13. Odegaard JI, Chawla A. Connecting type 1 and type 2 diabetes through innate immunity. Cold Spring Harbor perspectives in medicine. 2012;2(3):a007724.

14. Wu F, Zhao S, Yu B, Chen YM, Wang W, Song ZG, et al. Author Correction: A new coronavirus associated with human respiratory disease in China. Nature. 2020;580(7803):E7.

15. Guan WJ, Liang WH, He JX, Zhong NS. Cardiovascular comorbidity and its impact on patients with COVID-19. The European respiratory journal. 2020;55(6).

16. Li M, Dong Y, Wang H, Guo W, Zhou H, Zhang Z, et al. Cardiovascular disease potentially contributes to the progression and poor prognosis of COVID-19. Nutrition, metabolism, and cardiovascular diseases : NMCD. 2020;30(7):1061-7.

17. Emami A, Javanmardi F, Akbari A, Kojuri J, Bakhtiari H, Rezaei T, et al. Survival rate in hypertensive patients with COVID-19. Clinical and experimental hypertension. 2020:1-4.

18. Mohamed-Hussein A, Galal I, Mohamed Mmar, Ibrahim Meaa, Ahmed SB. Survival and 30-days hospital outcome in hospitalized COVID-19 patients in Upper Egypt: Multi-center study. medRxiv. 2020:2020.08.26.20180992.

19. Zhu L, She Z-G, Cheng X, Qin J-J, Zhang X-J, Cai J, et al. Association of Blood Glucose Control and Outcomes in Patients with COVID-19 and Pre-existing Type 2 Diabetes. Cell Metabolism. 2020;31(6):1068-77.e3.

20. Yan Y, Yang Y, Wang F, Ren H, Zhang S, Shi X, et al. Clinical characteristics and outcomes of patients with severe covid-19 with diabetes. BMJ open diabetes research & care. 2020;8(1).

21. Pranata R, Huang I, Lim MA, Wahjoepramono EJ, July J. Impact of cerebrovascular and cardiovascular diseases on mortality and severity of COVID-19-systematic review, meta-analysis, and meta-regression. Journal of stroke and cerebrovascular diseases : the official journal of National Stroke Association. 2020;29(8):104949.

22. Wang A-L, Zhong X, Hurd Y. Comorbidity and Sociodemographic determinants in COVID-19 Mortality in an US Urban Healthcare System. medRxiv. 2020:2020.06.11.20128926.

23. Oyelade T, Alqahtani J, Canciani G. Prognosis of COVID-19 in Patients with Liver and Kidney Diseases: An Early Systematic Review and Meta-Analysis. Tropical medicine and infectious disease. 2020;5(2).

